# PRICE COVID19 Data Report December 2021 *Pakistan Registry of Intensive Care*

**DOI:** 10.1101/2022.01.20.22269202

**Authors:** Ahmed Farooq, Aisha Kulsoom Mufti, Aneela Altaf, Arsalan Rahatullah, Arshad Taqi, Ashok Kumar, Attaur Rehman, Fakhir Raza Haidri, Iqbal Hussain, Irfan Malik, Jodat Saleem, Liaquat Ali, Mobin Chaudhry, Muhammad Sheharyar Ashraf, Mohiuddin Sheikh, Muhammad Ashraf Zia, Muhammad Asim, Muhammad Asim Rana, Muhammad Hayat, Muhammad Nasir Khoso, Naseem Ali Shaikh, Nawal Salahuddin, Qurat-ul-Ain Khan, Rana Imran Sikander, Rashid Nasim Khan, Saadiya Rizvi, Safdar Rehman, Sairah Babar, Syed Muneeb Ali, Abigail Beane, Aasiyah Rashan, Arjen M Dondorp, Chamira Kodippily, Himasha Muvindi, Dilanthi Priyadarshani, Ishara Udayanga, Rashan Haniffa, Sri Darshana, Srinivas Murthy, Thalha Rashan, Madiha Hashmi

## Abstract

Pakistan Registry of Intensive Care (PRICE) is a platform that has enabled standardized COVID-19 clinical data collection based on ISARIC/WHO Clinical Characterization Protocol. The near real-time data platform includes epidemiology, severity of illness, microbiology, treatment and outcomes of patients admitted with suspected or laboratory confirmed COVID19 infection to 67 intensive care and high dependency units across the country. Data has been extracted and analysed at regular intervals to inform stakeholders and improve care practices. This is our 28th report including all patients with suspected or confirmed COVID-19 from 26th March 2020 to 26th December 2021.

**Key findings** from 8624 patients who met eligibility criteria, are as follows:

⍰ Median age of 60 years (IQR 50-70).
⍰ The most common symptoms were shortness of breath (n = 6428, 77.8%), fever (n = 6091, 73.8%), and Cough (n = 3354, 38.9%)
⍰ The most common comorbidity was hypertension followed by diabetes.
⍰ During the course of illness 2804 (32.6%) patients received non-invasive ventilation, whereas 2474 (28.8%) patients had mechanical ventilation as their highest organ support. In addition, 2246 (26.1%) patients needed haemodynamic support and 1249 (14.7%) patients required renal replacement therapy as their highest organ support.
⍰ Median APACHE II score was 18
⍰ Overall mortality at ICU discharge was 39.2%
⍰ Increasing age and requirement for invasive mechanical ventilation were independent risk factors for mortality increased the risk of death

## Background

Coronavirus disease 2019 (COVID-19), is an ongoing pandemic of viral pneumonia.(1, 2) The first case of COVID-19 in Pakistan was confirmed from Karachi on February 26, 2020, and similar to other countries, Pakistan experienced an upsurge in COVID-19 infections.1287393 confirmed cases and 28784 deaths have been reported since the beginning of pandemic (3). There is paucity of clinical characterization data from low- and middle-income countries such as Pakistan for critically ill COVID-19 patients. (4-7). The Pakistan Registry of Intensive Care (PRICE), established in 2018, and part of the Wellcome funded “CRIT Care Asia” recruits more than 2000 monthly critical care admissions across a national network in Pakistan (8).

Aim of this report is to provide clinicians and stakeholders in critical care services in Pakistan with real-time information regarding the incidence, clinical character and service utilisation throughout the COVID-19 pandemic.

## Methods

The same PRICE platform was adapted to incorporate ISARIC/WHO Clinical Characterization Protocol (CCP) tiers 0-3 capturing clinical and epidemiological data of patients admitted to the collaborating high dependency (HDU) and intensive care units (ICU) with clinically suspected or laboratory confirmed COVID19 infection (9). A detailed description of data platform adaptation, data collection method and data management was published previously (9). Variables were added using a standardized nomenclature, Systematized Nomenclature of Medicine Clinical Terms (SNOMED CT), that was already operationalized in the platform, enhancing interoperability at the organizational level and facilitating sharing with ISARIC (10). Data is captured contemporaneously to clinical care via the digital platform, with 24 hr cadence. Inbuilt field validation and range checks promote data quality. Data completeness for ISARIC tiers 0 and 1 was above 97% during a recent external performance review (11).

### Data source

Information is entered in real-time from 67 participating units from 31 hospitals across the country and provides:

⍰ A notification when a suspected case of COVID-19 is admitted
⍰ Admission data for suspected COVID-19 cases, including demographics, characteristics, symptoms at presentation, comorbidities and travel history within 14 days
⍰ Severity of illness, severity of illness assessed by Acute Physiology And Chronic Health Evaluation II (APACHE II) score
⍰ Daily assessment of organ support and clinical treatments
⍰ Discharge information, including status and outcome of any investigations to confirm COVID-19 diagnosis.
⍰ All the enrolled patients in the registry were followed up to discharge and/or death.
⍰ Statistical analysis was performed using R software (R core team 2021).

The PRICE team are committed to ensuring the registry provides high quality information, whilst recognising the need for timely accessible information during these events. As such this information is collated and reported ‘as captured’. This information may undergo revision as analysis evolves and is disseminated for the purpose of primary stakeholders to support clinical care and resource allocation.

## Results

**This is the twenty-eighth report summarising the data on all suspected or confirmed COVID-19 cases from the 26th March 2020 to 26th December 2021 admitted to critical care units within the PRICE network that agreed to report data on severe acute respiratory infection (SARI)**.

### Number of Admissions

A total of 8624 critical care admissions with suspected or confirmed SARI were reported (Figure 1).

**Figure 1:**
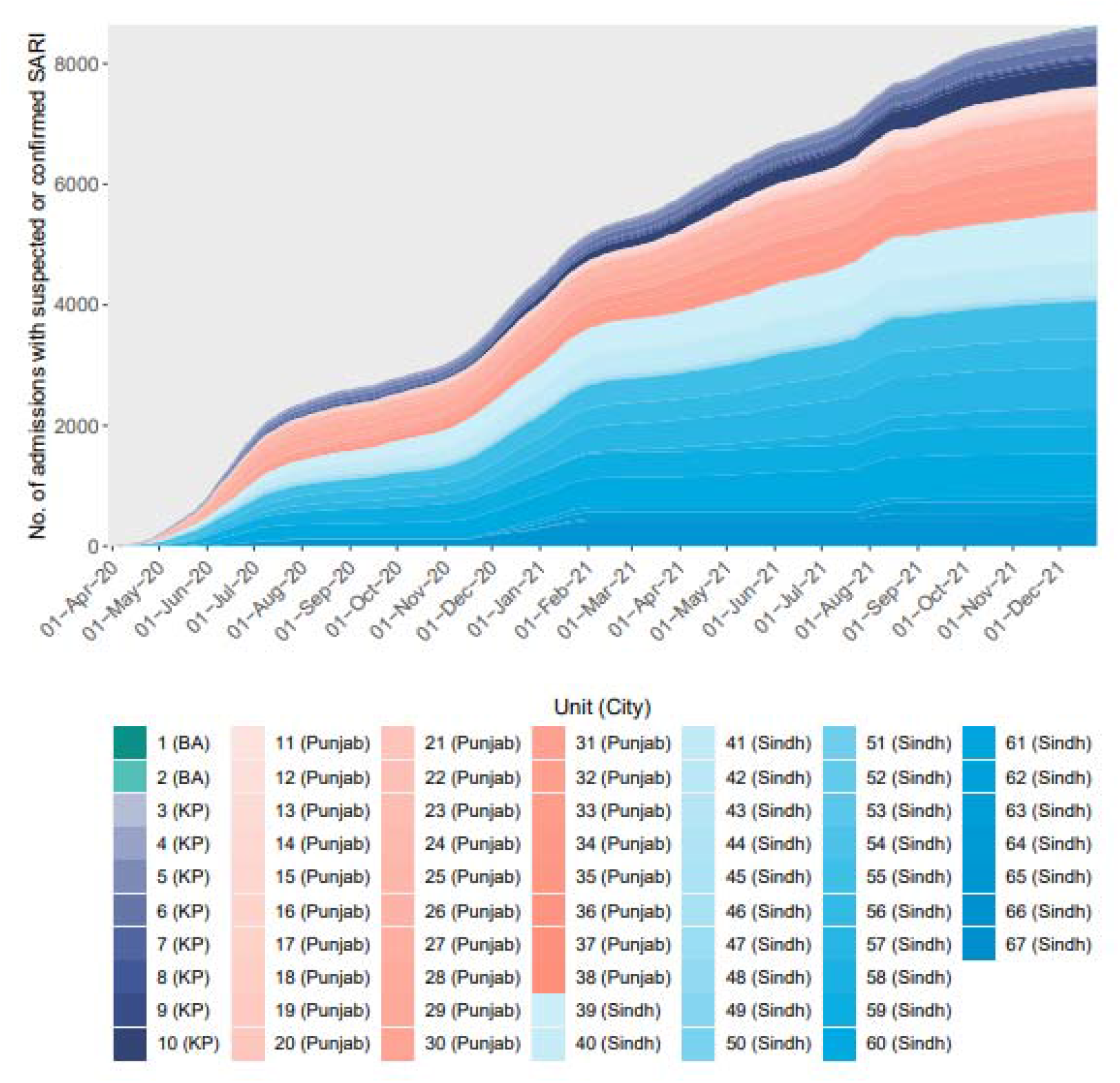
Number of ICU admissions with suspected or confirmed SARI by units.

### Characteristics of admitted patients

Characteristics of patients with suspected SARI admitted to participating critical care units are summarised in Table 1 and 2, and these are compared with the characteristics of critically ill patients with non-SARI viral pneumonia during 2018-2021 from the same registry. The most common presenting symptoms were shortness of breath, fever and cough with no sputum.

**Table 1:** Demographics and medical history of admitted patients.

| Demographics | Patients with suspected or confirmed SARI and admission data (N=8624) | Patients with viral pneumonia (non-SARI), 2018-2021 (N=153) |
| --- | --- | --- |
| **Age at admission (years)** |  |  |
| Mean (SD) | 58.6 (15.7) | 53.6 (19.6) |
| Median (IQR) | 60.0 (50.0, 70.0) | 56 (38.0, 68.0) |
| **Sex, n (%)** |  |  |
| Male | 5,384 (62.4) | 86 (56.2) |
| Female | 3,237 (37.5) | 67 (43.8) |
| Currently pregnant, n (females) [N=1560] | 87 (5.6) | * |
| **Ethnicity, n (%) [N=8200]** |  |  |
| Black | 3 (0.0) | * |
| East Asian | 10 (0.1) | * |
| Other | 2 (0.0) | * |
| South Asian | 8,178 (99.7) | * |
| West Asian | 6 (0.1) | * |
| White | 1 (0.0) | * |
| **Medical history** |  |  |
| **Comorbidities, n (%) [N=8624]** |  |  |
| At least one comorbidity present | 5,502 (63.8) | 66 (43.1) |
| Hypertension | 4,005 (46.4) | 52 (34.0) |
| Diabetes | 1,686 (19.6) | 42 (27.5) |
| Type 2 Diabetes | 1,033 (12.0) | 0 (0.0) |
| Cardiovascular diseases | 736 (8.5) | 3 (2.0) |
| Type 1 Diabetes | 493 (5.7) | 0 (0.0) |
| Other comorbidity | 260 (3.0) | 0 (0.0) |
| Asthma | 259 (3.0) | 1 (0.7) |
| Depression | 105 (1.2) | 0 (0.0) |
| Cerebrovascular disease | 96 (1.1) | 2 (1.3) |
| Hypothyroidism | 82 (1.0) | 0 (0.0) |
| Renal failure, Moderate to severe | 59 (0.7) | 1 (0.7) |
| Dementia | 54 (0.6) | 0 (0.0) |
| Respiratory disease, Severe moderate | 53 (0.6) | 1 (0.7) |
| Renal failure, Mild | 51 (0.6) | 2 (1.3) |
| Chronic pulmonary disease | 48 (0.6) | 0 (0.0) |
| Respiratory disease, Mild | 48 (0.6) | 0 (0.0) |
| CKD requiring dialysis | 47 (0.5) | 0 (0.0) |
| Angina | 37 (0.4) | 1 (0.7) |
| Tuberculosis | 29 (0.3) | 1 (0.7) |
| Valve disease | 23 (0.3) | 1 (0.7) |
| Obesity | 23 (0.3) | 0 (0.0) |
| Congestive heart failure | 22 (0.3) | 0 (0.0) |
| Other neurological condition | 18 (0.2) | 0 (0.0) |
| Hepatic disease, Moderate to severe | 18 (0.2) | 0 (0.0) |
| Hepatic disease, Mild | 16 (0.2) | 0 (0.0) |
| Type 1 diabetes with complications | 16 (0.2) | 0 (0.0) |
| Myocardial infarction | 15 (0.2) | 0 (0.0) |
| Hyperthyroidism | 15 (0.2) | 0 (0.0) |
| Epilepsy | 15 (0.2) | 0 (0.0) |
| Diabetes with complications | 15 (0.2) | 0 (0.0) |
| Rheumatological condition | 14 (0.2) | 0 (0.0) |
| Type 2 diabetes with complications | 10 (0.1) | 0 (0.0) |
| Tumor | 7 (0.1) | 0 (0.0) |
| Peripheral vascular disease | 7 (0.1) | 0 (0.0) |
| Hyperlipidemia | 7 (0.1) | 0 (0.0) |
| Inflammatory bowel | 6 (0.1) | 0 (0.0) |
| HIV | 5 (0.1) | 0 (0.0) |
| Interstitial lung disease | 4 (0.0) | 0 (0.0) |
| Peptic ulcer | 4 (0.0) | 0 (0.0) |
| Malignant neoplasm | 4 (0.0) | 0 (0.0) |
| Arrhythmia | 3 (0.0) | 0 (0.0) |
| Metastatic cancer | 2 (0.0) | 0 (0.0) |
| AIDS | 2 (0.0) | 0 (0.0) |
| Leukemia | 2 (0.0) | 0 (0.0) |
| Chronic hematologic disease | 2 (0.0) | 0 (0.0) |
| GI bleeding | 1 (0.0) | 0 (0.0) |
| Lymphoma | 1 (0.0) | 0 (0.0) |
| Primary lung cancer | 1 (0.0) | 0 (0.0) |
| Non diabetic endocrine | 1 (0.0) | 0 (0.0) |
| Malnutrition | 1 (0.0) | 0 (0.0) |

<sup>1</sup> \*Data not available prior to SARI data collection.
<sup>2</sup> Includes patients with confirmed viral pneumonia (n=274) and those with pneumonia (other, but not bacterial or fungal) (n=99)

**Table 2:**
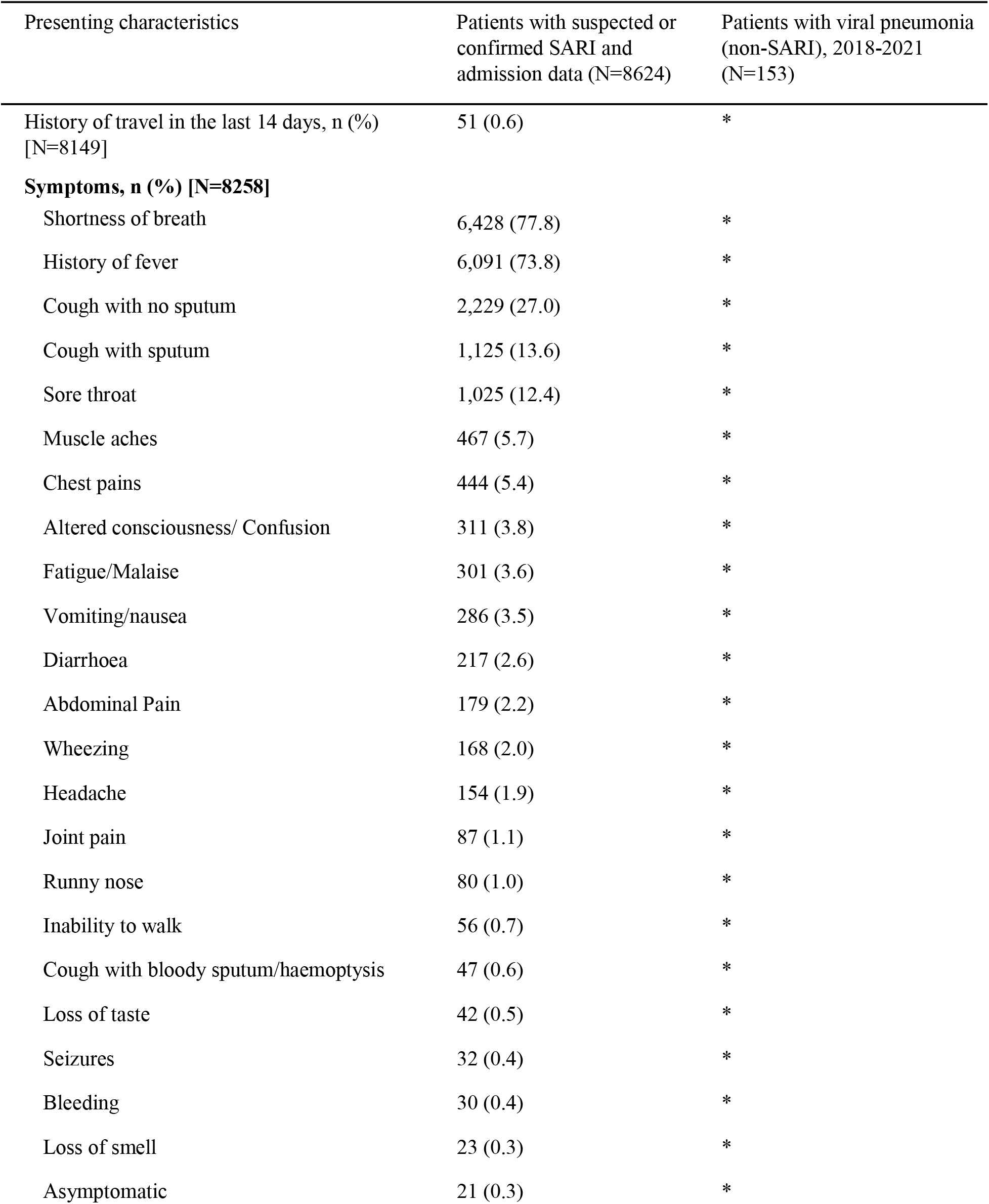
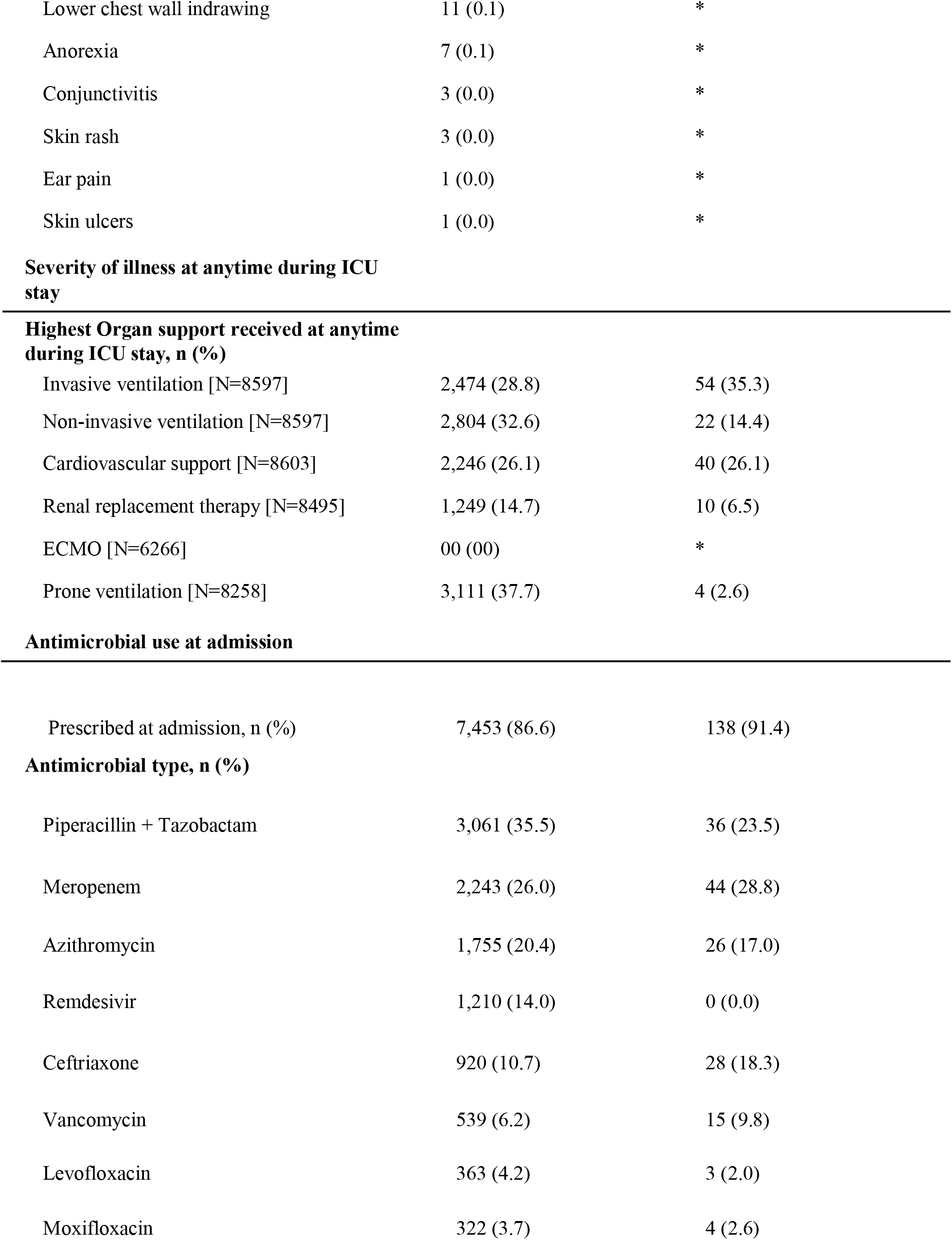
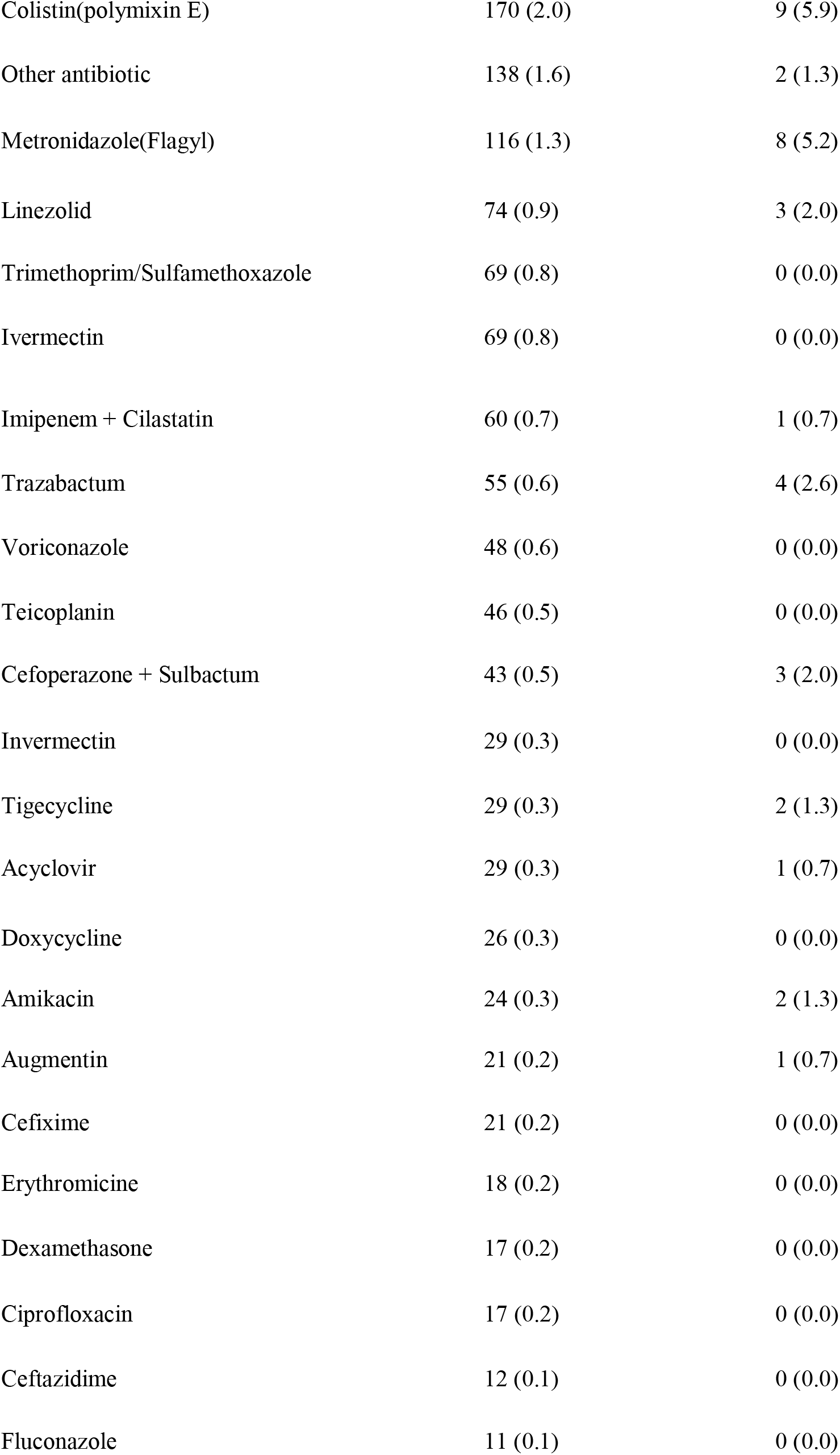
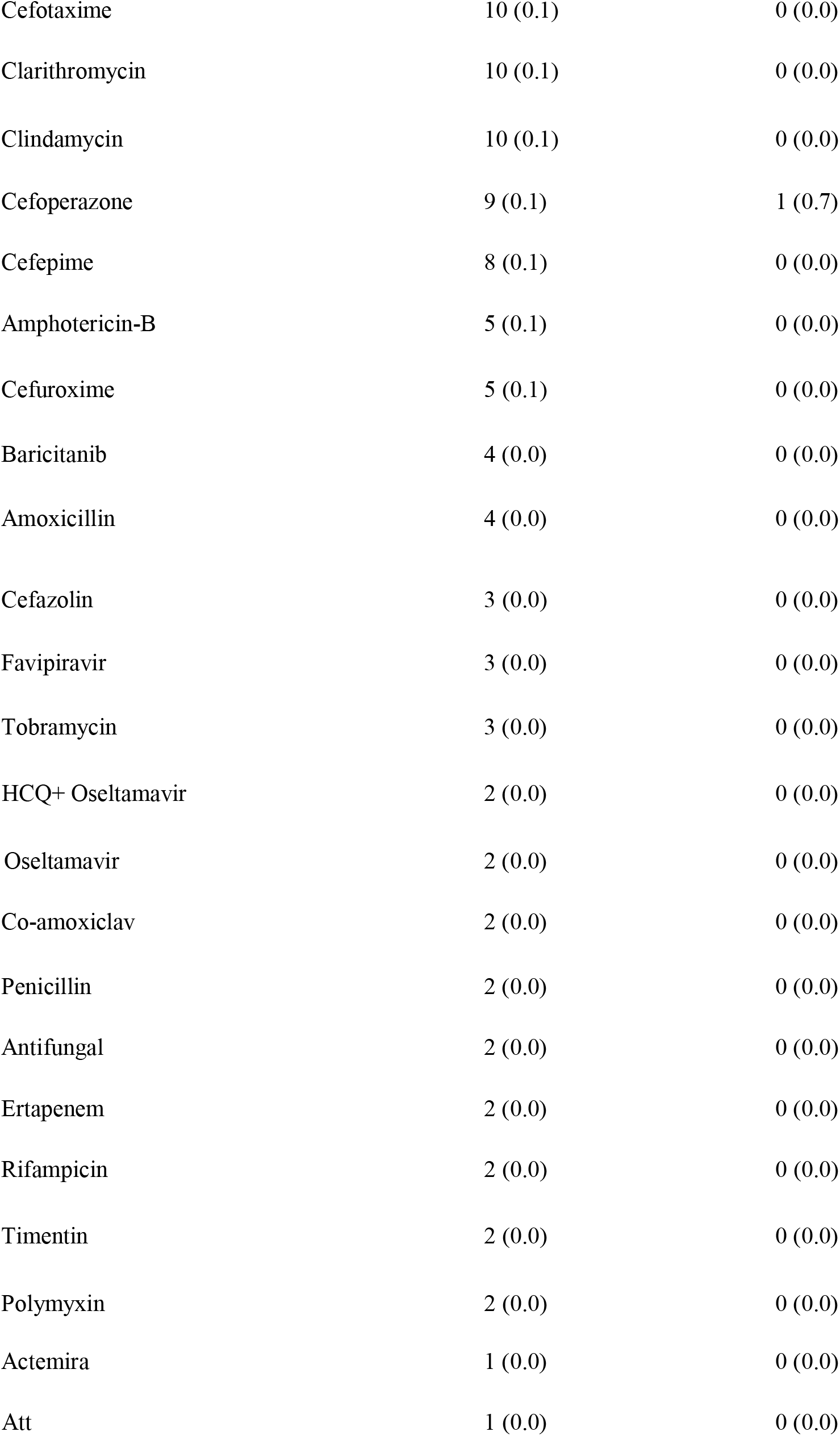
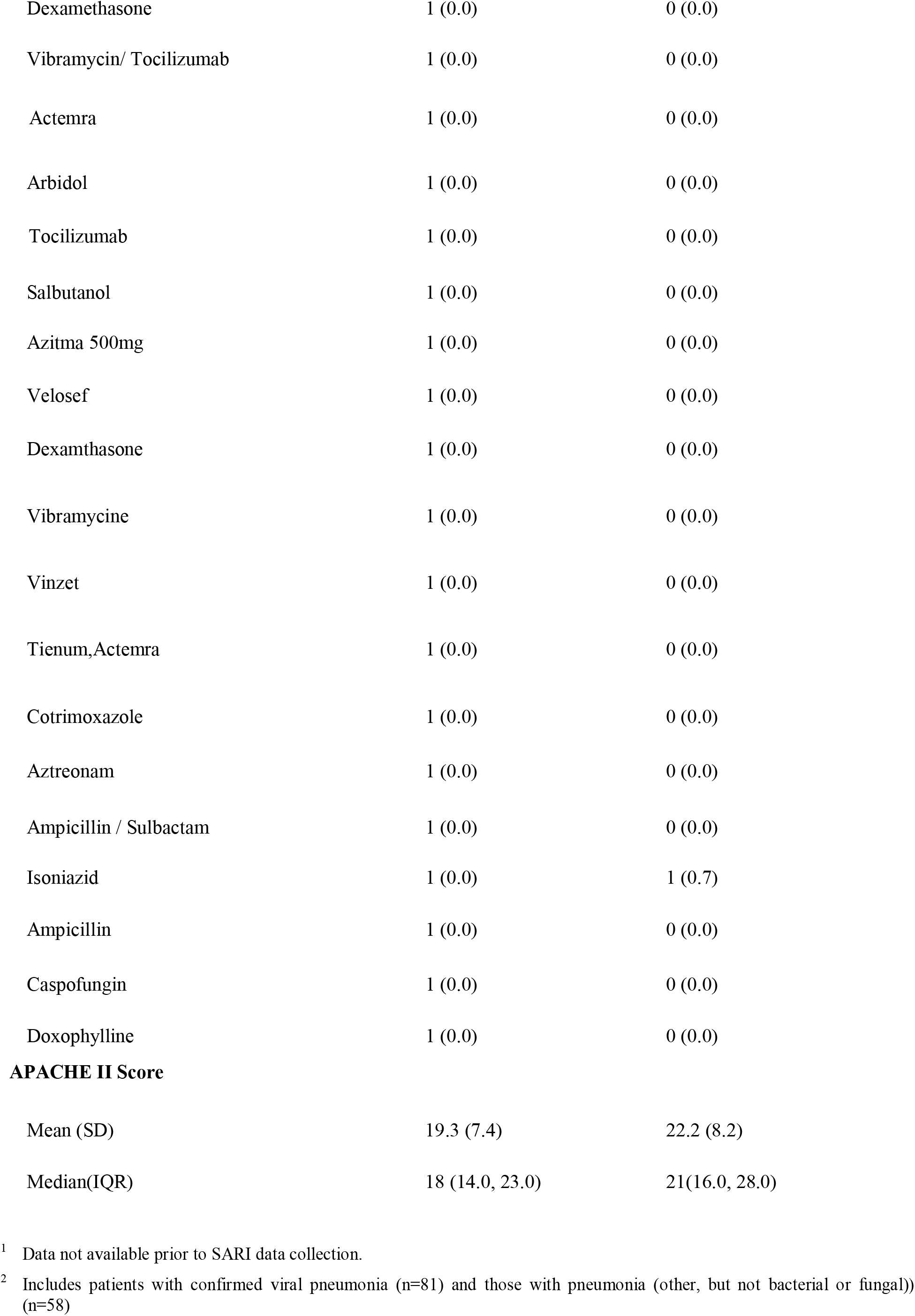
Clinical characteristics on admission

| Presenting characteristics | Patients with suspected or confirmed SARI and admission data (N=8624) | Patients with viral pneumonia (non-SARI), 2018-2021 (N=153) |
| --- | --- | --- |
| History of travel in the last 14 days, n (%) [N=8149] | 51 (0.6) | * |
| **Symptoms, n (%) [N=8258]** |  |  |
| Shortness of breath | 6,428 (77.8) | * |
| History of fever | 6,091 (73.8) | * |
| Cough with no sputum | 2,229 (27.0) | * |
| Cough with sputum | 1,125 (13.6) | * |
| Sore throat | 1,025 (12.4) | * |
| Muscle aches | 467 (5.7) | * |
| Chest pains | 444 (5.4) | * |
| Altered consciousness/ Confusion | 311 (3.8) | * |
| Fatigue/Malaise | 301 (3.6) | * |
| Vomiting/nausea | 286 (3.5) | * |
| Diarrhoea | 217 (2.6) | * |
| Abdominal Pain | 179 (2.2) | * |
| Wheezing | 168 (2.0) | * |
| Headache | 154 (1.9) | * |
| Joint pain | 87 (1.1) | * |
| Runny nose | 80 (1.0) | * |
| Inability to walk | 56 (0.7) | * |
| Cough with bloody sputum/haemoptysis | 47 (0.6) | * |
| Loss of taste | 42 (0.5) | * |
| Seizures | 32 (0.4) | * |
| Bleeding | 30 (0.4) | * |
| Loss of smell | 23 (0.3) | * |
| Asymptomatic | 21 (0.3) | * |
| Lower chest wall indrawing | 11 (0.1) | * |
| Anorexia | 7 (0.1) | * |
| Conjunctivitis | 3 (0.0) | * |
| Skin rash | 3 (0.0) | * |
| Ear pain | 1 (0.0) | * |
| Skin ulcers | 1 (0.0) | * |
| Highest Organ support received at anytime during ICU stay, n (%) |  |  |
| Invasive ventilation [N=8597] | 2,474 (28.8) | 54 (35.3) |
| Non-invasive ventilation [N=8597] | 2,804 (32.6) | 22 (14.4) |
| Cardiovascular support [N=8603] | 2,246 (26.1) | 40 (26.1) |
| Renal replacement therapy [N=8495] | 1,249 (14.7) | 10 (6.5) |
| ECMO [N=6266] | 00 (00) | * |
| Prone ventilation [N=8258] | 3,111 (37.7) | 4 (2.6) |
| Antimicrobial use at admission |  |  |
| Prescribed at admission, n (%) | 7,453 (86.6) | 138 (91.4) |
| Antimicrobial type, n (%) |  |  |
| Piperacillin + Tazobactam | 3,061 (35.5) | 36 (23.5) |
| Meropenem | 2,243 (26.0) | 44 (28.8) |
| Azithromycin | 1,755 (20.4) | 26 (17.0) |
| Remdesivir | 1,210 (14.0) | 0 (0.0) |
| Ceftriaxone | 920 (10.7) | 28 (18.3) |
| Vancomycin | 539 (6.2) | 15 (9.8) |
| Levofloxacin | 363 (4.2) | 3 (2.0) |
| Moxifloxacin | 322 (3.7) | 4 (2.6) |
| Colistin(polymixin E) | 170 (2.0) | 9 (5.9) |
| Other antibiotic | 138 (1.6) | 2 (1.3) |
| Metronidazole(Flagyl) | 116 (1.3) | 8 (5.2) |
| Linezolid | 74 (0.9) | 3 (2.0) |
| Trimethoprim/Sulfamethoxazole | 69 (0.8) | 0 (0.0) |
| Ivermectin | 69 (0.8) | 0 (0.0) |
| Imipenem + Cilastatin | 60 (0.7) | 1 (0.7) |
| Trazabactam | 55 (0.6) | 4 (2.6) |
| Voriconazole | 48 (0.6) | 0 (0.0) |
| Teicoplanin | 46 (0.5) | 0 (0.0) |
| Cefoperazone + Sulbactam | 43 (0.5) | 3 (2.0) |
| Invermectin | 29 (0.3) | 0 (0.0) |
| Tigecycline | 29 (0.3) | 2 (1.3) |
| Acyclovir | 29 (0.3) | 1 (0.7) |
| Doxycycline | 26 (0.3) | 0 (0.0) |
| Amikacin | 24 (0.3) | 2 (1.3) |
| Augmentin | 21 (0.2) | 1 (0.7) |
| Cefixime | 21 (0.2) | 0 (0.0) |
| Erythromicine | 18 (0.2) | 0 (0.0) |
| Dexamethasone | 17 (0.2) | 0 (0.0) |
| Ciprofloxacin | 17 (0.2) | 0 (0.0) |
| Ceftazidime | 12 (0.1) | 0 (0.0) |
| Fluconazole | 11 (0.1) | 0 (0.0) |
| Cefotaxime | 10 (0.1) | 0 (0.0) |
| Clarithromycin | 10 (0.1) | 0 (0.0) |
| Clindamycin | 10 (0.1) | 0 (0.0) |
| Cefoperazone | 9 (0.1) | 1 (0.7) |
| Cefepime | 8 (0.1) | 0 (0.0) |
| Amphotericin-B | 5 (0.1) | 0 (0.0) |
| Cefuroxime | 5 (0.1) | 0 (0.0) |
| Baricitanib | 4 (0.0) | 0 (0.0) |
| Amoxicillin | 4 (0.0) | 0 (0.0) |
| Cefazolin | 3 (0.0) | 0 (0.0) |
| Favipiravir | 3 (0.0) | 0 (0.0) |
| Tobramycin | 3 (0.0) | 0 (0.0) |
| HCQ+ Oseltamavir | 2 (0.0) | 0 (0.0) |
| Oseltamavir | 2 (0.0) | 0 (0.0) |
| Co-amoxiclav | 2 (0.0) | 0 (0.0) |
| Penicillin | 2 (0.0) | 0 (0.0) |
| Antifungal | 2 (0.0) | 0 (0.0) |
| Ertapenem | 2 (0.0) | 0 (0.0) |
| Rifampicin | 2 (0.0) | 0 (0.0) |
| Timentin | 2 (0.0) | 0 (0.0) |
| Polymyxin | 2 (0.0) | 0 (0.0) |
| Actemira | 1 (0.0) | 0 (0.0) |
| Att | 1 (0.0) | 0 (0.0) |
| Dexamethasone | 1 (0.0) | 0 (0.0) |
| Vibramycin/ Tocilizumab | 1 (0.0) | 0 (0.0) |
| Actemra | 1 (0.0) | 0 (0.0) |
| Arbidol | 1 (0.0) | 0 (0.0) |
| Tocilizumab | 1 (0.0) | 0 (0.0) |
| Salbutanol | 1 (0.0) | 0 (0.0) |
| Azitma 500mg | 1 (0.0) | 0 (0.0) |
| Velosef | 1 (0.0) | 0 (0.0) |
| Dexamthasone | 1 (0.0) | 0 (0.0) |
| Vibramycine | 1 (0.0) | 0 (0.0) |
| Vinzet | 1 (0.0) | 0 (0.0) |
| Tienum,Actemra | 1 (0.0) | 0 (0.0) |
| Cotrimoxazole | 1 (0.0) | 0 (0.0) |
| Aztreonam | 1 (0.0) | 0 (0.0) |
| Ampicillin / Sulbactam | 1 (0.0) | 0 (0.0) |
| Isoniazid | 1 (0.0) | 1 (0.7) |
| Ampicillin | 1 (0.0) | 0 (0.0) |
| Caspofungin | 1 (0.0) | 0 (0.0) |
| Doxophylline | 1 (0.0) | 0 (0.0) |
| **APACHE II Score** |  |  |
| Mean (SD) | 19.3 (7.4) | 22.2 (8.2) |
| Median(IQR) | 18 (14.0, 23.0) | 21(16.0, 28.0) |

<sup>1</sup> Data not available prior to SARI data collection.
<sup>2</sup> Includes patients with confirmed viral pneumonia (n=81) and those with pneumonia (other, but not bacterial or fungal)) (n=58)

### Care provided in critical care and discharge status

Discharge data was available for 8521 patients. There were 5183 patients discharged alive from critical care (Figure 2). Table 3 outlines the length of stay and outcome for critical care admissions, and compares it with non-SARI viral pneumonia patients during 2018-2021.

**Figure 2:**
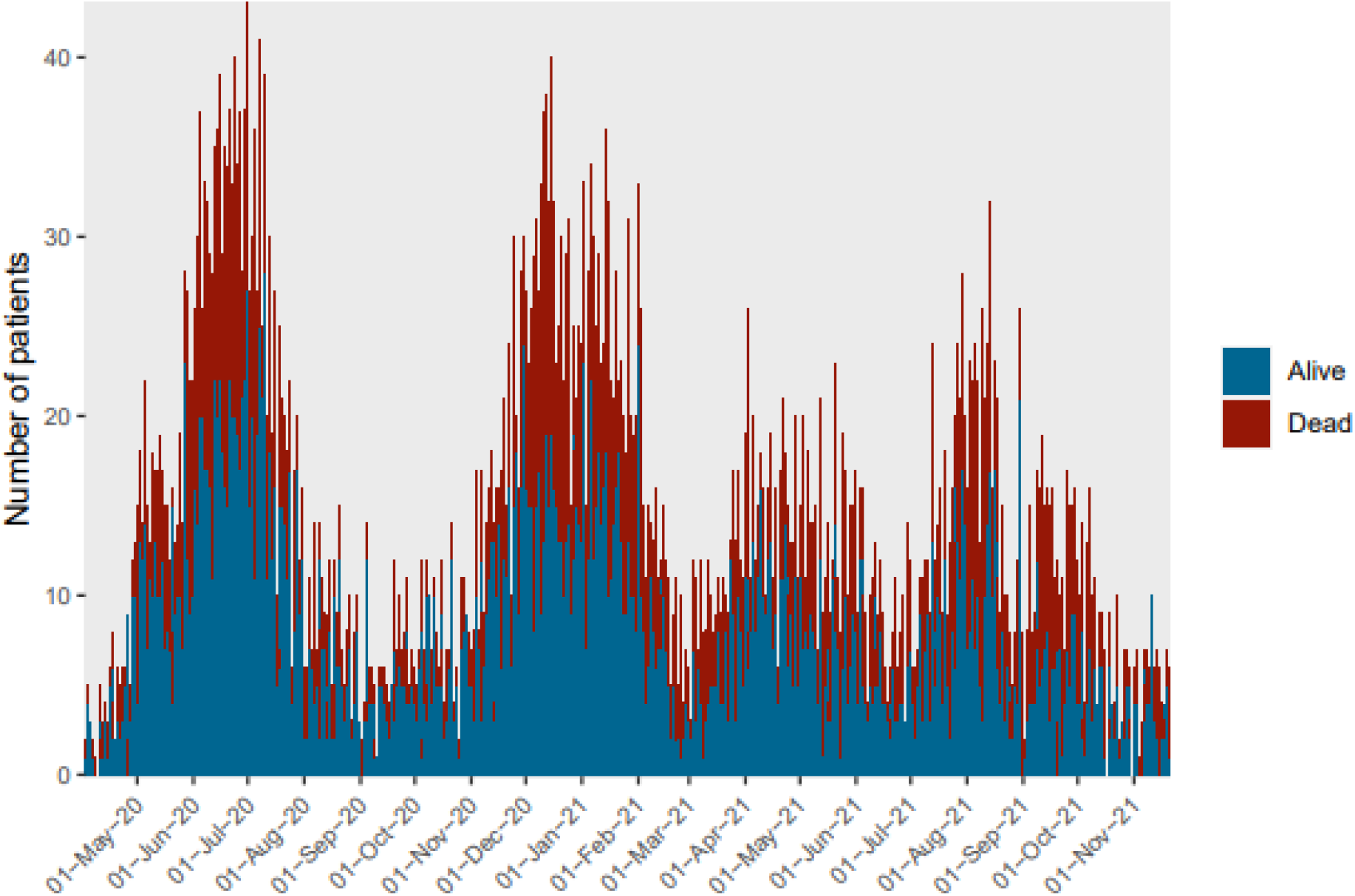
Patient outcomes at ICU discharge

**Table 3:** Outcome and length of stay for patients admitted to critical care with suspected or confirmed SARI

| Critical care unit outcome | Patients with suspected or confirmed SARI and discharge data (N=8521) | Patients with viral pneumonia (non-SARI), 2018-2021 (N=153) |
| --- | --- | --- |
| COVID-19 confirmed at discharge, n (%) [N=7579] | 6234 (82.3) | * |
| **Outcome at ICU discharge, n (%) [N=8521]** |  |  |
| Alive | 5,183 (60.8) | 107 (69.9) |
| Deceased | 3,338 (39.2) | 46 (30.1) |
| **Outcome for patients invasively ventilated at admission, n (%) [N=1560]** |  |  |
| Alive | 421 (27.0) | 23 (47.9) |
| Deceased | 1,139 (73.0) | 25 (52.1) |
| **Outcome for patient non-invasively ventilated at admission, n (%) [N=1742]** |  |  |
| Alive | 899 (51.6) | 6 (60.0) |
| Deceased | 843 (48.4) | 4 (40.0) |
| **Outcome for vaccinated patients (at least one dose received), n (%) [N=298]\*\*** |  |  |
| Alive | 198 (66.4) | * |
| Deceased | 100 (33.6) | * |
| **Outcome for non-vaccinated patients, n (%) [N=479]** |  |  |
| Alive | 300 (62.6) | * |
| Deceased | 179 (37.4) | * |
| **Length of stay in critical care (days), median (IQR)** |  |  |
| Alive | 3.4 (1.7, 6.1) | 4.0 (1.9, 6.8) |
| Deceased | 4.0 (1.6, 8.0) | 2.5 (0.8, 5.9) |

<sup>1</sup> \*Data not available prior to SARI data collection.
<sup>2</sup> \*\*Vaccine information was enabled in the registry from 9th July 2021.
<sup>3</sup> Includes patients with confirmed viral pneumonia (n=81) and those with pneumonia (other, but not bacterial or fungal)) (n=58)

### COVID-19 vaccine information

(Capturing COVID-19 vaccine information in the registry was enabled from 9th July 2021)

One thousand six hundred seventy-five critical care admissions of suspected or confirmed SARI have been notified from 9th July 2021 to 26th December 2021. Of them, COVID-19 vaccine information has been received for 841(50.2%) patients. There, 321 (38.2%) patients were vaccinated while 520 (61.8%) patients have not got any COVID-19 vaccine.

**Table 5:** COVID-19 vaccine information of the patients admitted to critical care with suspected or confirmed SARI

| Vaccine status | Count [ N = 841] |
| --- | --- |
| **Vaccinated, n(%)** | 321 (38.2) |
| First dose received | 321 (100.0) |
| Second dose received | 151 (47.0) |
| Booster dose received | 8 (2.5) |
| **Non vaccinated, n(%)** | 520 (61.8) |

Of 321 vaccinated patients, all have received their first dose, while 151 (47%) have received their second dose. 8 patients (2.5%) have received their booster dose.

## Data Availability

All data produced in the present work are contained in the manuscript

## Acknowledgement

The authors would like to acknowledge all the collaborators of PRICE and the Pakistan Institute of Living and Learning.

